# SARS-CoV-2 Omicron triggers cross-reactive neutralization and Fc effector functions in previously vaccinated, but not unvaccinated individuals

**DOI:** 10.1101/2022.02.10.22270789

**Authors:** Simone I. Richardson, Vimbai Sharon Madzorera, Holly Spencer, Nelia P. Manamela, Mieke A. van der Mescht, Bronwen E. Lambson, Brent Oosthuysen, Frances Ayres, Zanele Makhado, Thandeka Moyo-Gwete, Nonkululeko Mzindle, Thopisang Motlou, Amy Strydom, Adriano Mendes, Houriiyah Tegally, Zelda de Beer, Talita Roma de Villiers, Annie Bodenstein, Gretha van den Berg, Marietjie Venter, Tulio de Oliviera, Veronica Ueckermann, Theresa M. Rossouw, Michael T. Boswell, Penny L. Moore

## Abstract

The SARS-CoV-2 Omicron variant escapes neutralizing antibodies elicited by vaccines or infection. However, whether Omicron triggers cross-reactive humoral responses to other variants of concern (VOCs) remains unknown. We use plasma from 20 unvaccinated and seven vaccinated individuals infected by Omicron BA.1 to test binding, Fc effector function and neutralization against VOCs. In unvaccinated individuals, Fc effector function and binding antibodies target Omicron and other VOCs at comparable levels. However, Omicron BA.1-triggered neutralization is not extensively cross-reactive for VOCs (14 to 31-fold titer reduction) and we observe 4-fold decreased titers against Omicron BA.2. In contrast, vaccination followed by breakthrough Omicron infection was associated with improved cross-neutralization of VOCs, with titers exceeding 1:2,100. This has important implications for vulnerability of unvaccinated Omicron-infected individuals to reinfection by circulating and emerging VOCs. While Omicron-based immunogens may be adequate boosters, they are unlikely to be superior to existing vaccines for priming in SARS-CoV-2 naïve individuals.

## Introduction

The emergence of the SARS-CoV-2 Omicron (B.1.1.529) variant of concern (VOC) in November 2021 coincided with the fourth wave of South Africa’s COVID-19 epidemic (Viana et al., 2022). Omicron is defined by multiple mutations across its genome, including more than 30 mutations in spike, many of which are associated with immune evasion (Viana et al., 2022). Omicron has further evolved in additional sub-lineages, including BA.1 and BA.2, with 17 amino acid, 3 deletions and one insertion distinguishing the two sub-lineages, many in the N terminal domain (NTD) and receptor binding domain (RBD). These mutations confer neutralization escape in vaccinees and in donors previously infected with other variants (Cele et al., 2021; Garcia-Beltran et al., 2022; Schmidt et al., 2021; Sievers et al., 2022).

In contrast to neutralization, antibody binding to the Omicron variant is preserved, as observed for other VOCs (Carreño et al., 2021; Wibmer et al., 2021). Although Fc receptor binding was substantially reduced in antibodies tested against Omicron (Bartsch et al., 2021), functional Fc effector responses have not yet been reported. The ability of antibodies to bind the Omicron spike suggests that cytotoxic effector functions driven by antibodies may also be retained, as for earlier VOCs (Kaplonek et al., 2022; Richardson et al., 2022). This, along with the finding that T cells triggered by infection or vaccination are cross-reactive for Omicron (Gao et al., 2022; Keeton et al., 2022; Tarke et al., 2022), likely contributes to maintained vaccine effectiveness against severe disease following Omicron infection (Collie et al., 2022).

While there is substantial data showing that Omicron evades neutralizing] responses, little is known about the humoral response that Omicron infection itself triggers. Defining the capacity of Omicron to trigger cross-reactive binding, neutralizing and Fc effector responses antibodies will inform its potential to protect from reinfection by currently circulating or emerging VOCs. This is of particular relevance for developing countries such as South Africa, where vaccination levels are low and genomic surveillance indicates continued circulation of Delta and other variants. Furthermore, little is known about the activity of antibodies triggered by BA.1 infection against BA.2 (Yamasoba et al., 2022; Yu et al., 2022). Secondly, these data inform the potential immunogenicity of Omicron-based vaccines under development by several companies.

## Results

Plasma from individuals infected in the fourth wave of COVID-19 pandemic in South Africa were used to assess cross-reactivity against different VOCs for binding, Fc effector function and neutralization. Plasma was used from 27 hospitalized individuals from Tshwane District Hospital recruited between 25 November 2021 and 20 December 2021 when Omicron BA.1 was responsible for >90% of infections (Viana et al., 2022) (Table S1). Of these, seven plasma samples had matched nasal swabs, and all were Omicron BA.1 infections by sequencing. Twenty individuals were unvaccinated with no history of previous symptomatic COVID-19 infection. Seven individuals had previously been vaccinated with either one dose of Ad26.CoV2.S (n=2) or two doses of BNT162b2 (n=5) at least 56 days (56-163 days) prior to infection. Samples were taken a median of four days (1-10 days) after a positive PCR test. The median ages of the vaccinated and unvaccinated individuals were similar (58 and 64 respectively), and infections ranged from mild to severe as determined by WHO scoring (Table S1).

We first compared binding antibody levels, as measured by enzyme-linked immunosorbent assay (ELISA) against the ancestral D614G, Beta, Delta and Omicron BA.1 spikes. In unvaccinated individuals, binding antibody titers against Omicron BA.1 were highest, as expected, and were detectable in all donors. Although we observed statistically significant 2.2, 1.8 and 1.7-fold decreases in binding to D614G, Beta and Delta respectively in this group, Omicron BA.1-triggered antibodies were fairly cross-reactive for all variants tested, losing activity against other VOCs in 10-25% of individuals (Figure 1A, C). In previously vaccinated individuals who experienced breakthrough infection with Omicron BA.1, binding was higher against Omicron BA.1 than in unvaccinated individuals (geometric mean titer (GMT) of 2.96 versus 1.95) (Figure 1B, C). Furthermore, antibodies from these vaccinated individuals exhibited higher levels of cross-reactivity against all variants, and no significant fold losses were observed (Figure 1B).

**Figure 1:**
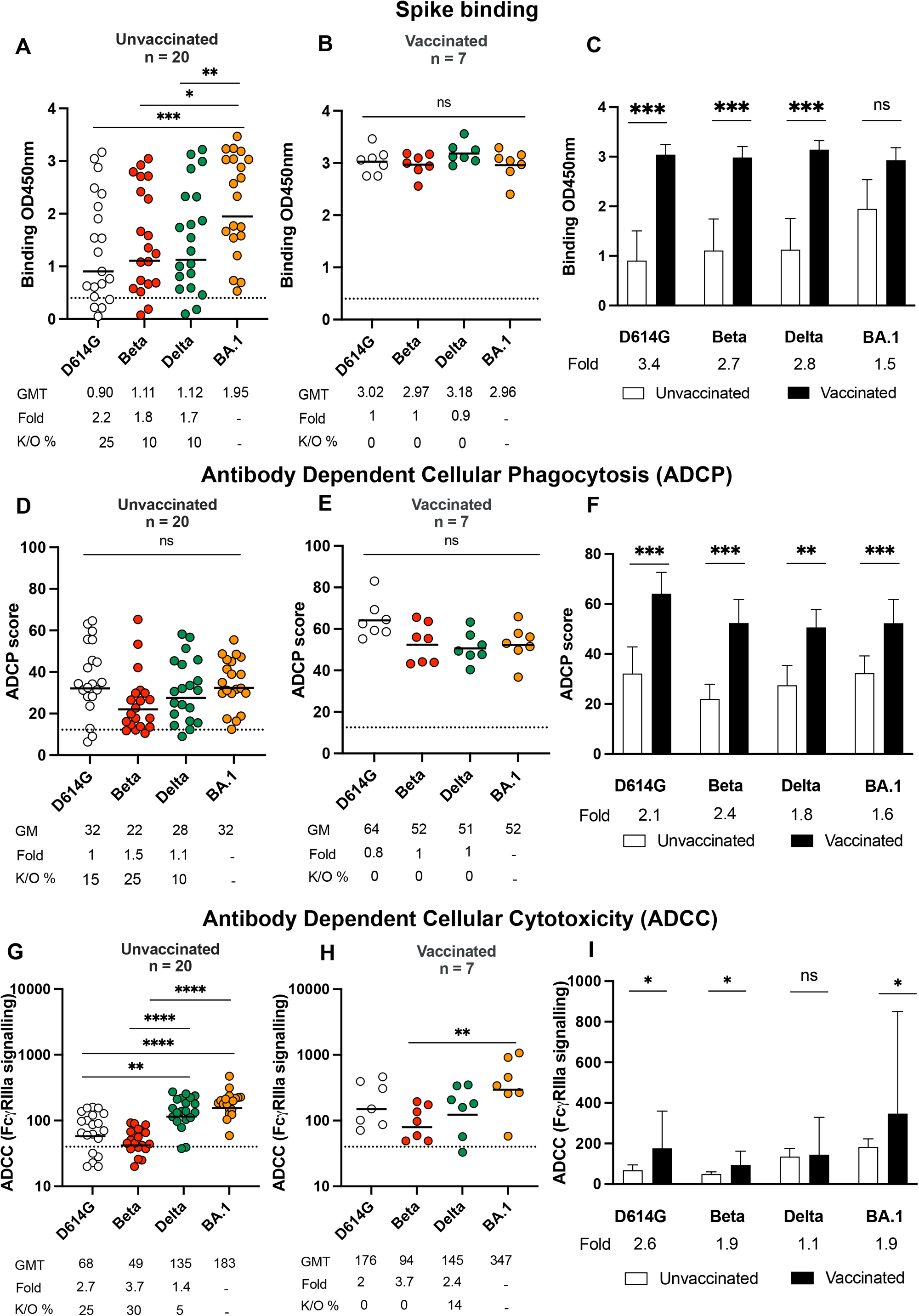
Binding and Fc effector function elicited by Omicron infection is cross-reactive against several variants of concern. Antibody binding measured by ELISA in (A) unvaccinated individuals (n=20) or (B) individuals vaccinated with either one dose of Ad26.CoV2.S or two doses of BNT162b2 (n=7) and infected by Omicron against D614G, Beta, Delta and Omicron BA.1 spike proteins. (C) Bars show geometric mean binding titers for vaccinated (black) and unvaccinated (white) individuals against variants of concern. Antibody-dependent cellular phagocytosis (ADCP) of (D) unvaccinated and (E) vaccinated individuals is represented as the percentage of monocytic cells that take up spike coated beads (D614G, Beta, Delta and Omicron BA.1) multiplied by their geometric mean fluorescence intensity (MFI). (F) Bars show geometric mean ADCP scores for vaccinated (black) and unvaccinated (white) individuals against variants of concern. Antibody dependent cellular cytotoxicity (ADCC) in (G) unvaccinated and (H) vaccinated individuals shown as relative light units (RLU) signaling through FcγRIIIa expressing cells. (I) Bars show geometric mean activity for vaccinated (black) and unvaccinated (white) individuals against variants of concern. All data are representative of two independent experiments. For dot plots, lines indicate geometric mean titer (GMT) also represented below the plot with fold decrease and knock-out (K/O) of activity for other variants as a percentage relative to Omicron BA.1. Dotted lines indicate the limit of detection of the particular assay. For bar charts, bars indicate median of function, with error bars showing standard deviations with fold decreases relative to vaccinated individuals indicated below the plot. Statistical significance across variants is shown by Friedman test with Dunn’s correction and between vaccinated and unvaccinated samples by the Mann Whitney test. *p<0.05; **p<0.01; ***p<0.001; ****p<0.0001 and ns = non-significant.

As spike binding antibodies perform Fc effector functions known to contribute to reduced disease severity and vaccine efficacy (Yu et al., 2020; Zohar et al., 2020), we examined antibody dependent cellular phagocytosis (ADCP) and antibody dependent cellular cytotoxicity (ADCC) in both groups. For unvaccinated individuals, ADCP against Omicron BA.1 was detected in all 20 individuals with a geometric mean (GM) score of 32 (Figure 1D). Against VOCs, we observed less than two-fold reduction in activity across variants with 15%, 25% and 10% of individuals losing ADCP activity against D614G, and Beta and Delta respectively. For vaccinated individuals, non-significant reductions against D614G, Beta and Delta were observed relative to Omicron BA.1 and all donors exhibited activity against the panel of VOC tested here (Figure 1E). Compared to unvaccinated individuals, significantly higher levels of ADCP were observed in vaccinated individuals infected with Omicron BA.1, mirroring the binding antibodies (Figure 1E, F).

In contrast to binding and ADCP, ADCC in unvaccinated individuals showed significant losses against D614G (3-fold loss) and Beta (4-fold loss). However, like ADCP and binding antibodies, ADCC activity against Delta was retained (Figure 1G). In this group, Omicron BA.1-triggered ADCC was undetectable against D614G and Beta in 25% and 30% of plasma samples, respectively. After previous vaccination, Omicron BA.1 breakthrough infections resulted in overall preservation of ADCC against VOCs, with only one individual showing undetectable activity against Delta (Figure 1H). Levels of ADCC in previously vaccinated donors were significantly higher than those in unvaccinated individuals, except for Delta, where similar ADCC activity was observed in both groups (Figure 1I).

Lastly, we measured neutralizing antibody responses to Omicron BA.1, and assessed cross-reactivity for VOCs. In addition to the variants used above, we also tested C.1.2, a variant with several neutralization evasive mutations, which circulated at low levels during the third and fourth COVID-19 waves in South Africa (Scheepers et al., 2021) and BA.2, a sub-lineage of Omicron. Against Omicron BA.1, unvaccinated individuals showed potent neutralization, with a GMT of 3,043. Neutralization of BA.2 was 4-fold lower than against BA.1, with a GMT of 826, and only 5% samples showed knockout of neutralization. However, against VOCs, we saw dramatic reductions of 19-, 31-, 16- and 14-fold in titer for D614G, Beta, C.1.2 and Delta, respectively, and knock out ranging from 27% to 45% of plasma tested (Figure 2A), particularly evident for Beta. This preferential neutralization of Omicron (the infecting strain) over other VOCs is in sharp contrast to samples from the previous three waves of infection in South Africa (for which the infecting virus was D614G, Beta or Delta, respectively) where neutralizing activity against D614G was substantially higher than against Omicron BA.1 (Figure S1A and B). Similarly, samples from BNT162b2 or Ad26.COV2.S showed higher activity against D614G than Omicron (Supplementary Figure 1A and B), consistent with previous studies (Cele et al., 2021; Garcia-Beltran et al., 2022).

**Figure 2:**
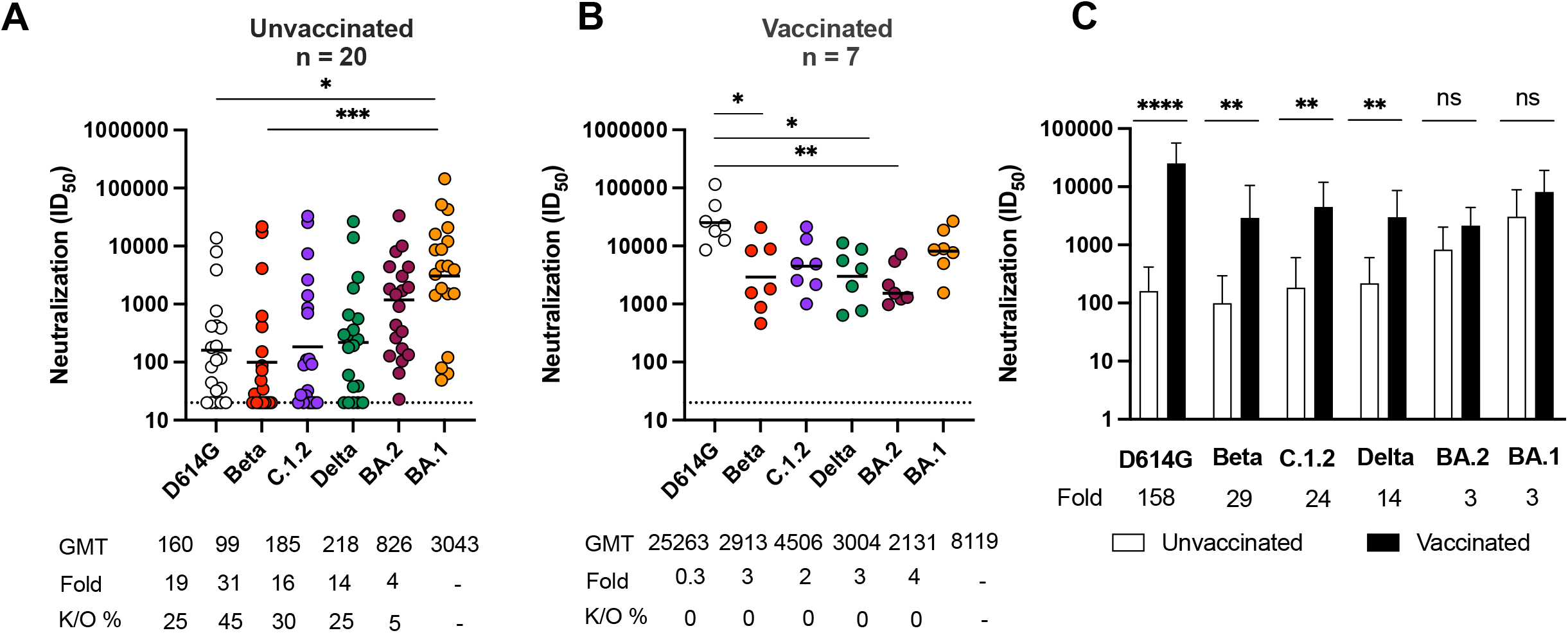
Omicron triggers cross-variant neutralizing antibodies which are broadened by vaccination. Neutralization titer (ID_50_) of Omicron-infected plasma against D614G, Beta, C.1.2, Delta, Omicron BA.2 and Omicron BA.1 pseudoviruses shown for (A) unvaccinated individuals (n=20) or (B) individuals vaccinated with either one dose of Ad26.CoV.2S or two doses of BNT162b2 (n=7). Lines indicate geometric mean titer (GMT) also represented below the plot with fold decrease and knock-out (K/O) of activity for other variants as a percentage relative to Omicron BA.1. Dotted lines indicate the limit of detection of the assay. Statistical significance across variants is shown by Friedman test with Dunns correction. (C) Bars show geometric mean neutralization titers for vaccinated (black) and unvaccinated (white) individuals against variants of concern with error bars showing standard deviations with fold decreases relative to vaccinated individuals indicated below the plot. Statistical significance between vaccinated and unvaccinated samples by the Mann Whitney test. *p<0.05; **p<0.01; ***p<0.001; ****p<0.0001 and ns = non-significant. All data are representative of two independent experiments.

This loss of neutralization was mitigated by prior vaccination, with all seven breakthrough infections resulting in a significantly increased GMT of 8,119 against Omicron BA.1, GMT of 2,131 against Omicron BA.2, and high titers against all VOCs (25,263 for D614G; 2,913 for Beta; 4,506 against C.1.2 and 3004 against Delta) (Figure 2B, C). These greatly enhanced titers resulted in far greater fold increases between previously vaccinated individuals and unvaccinated individuals (158, 29,, 24, 14, 3 and 3-fold for D614G, Beta, C.1.2, Delta, Omicron BA.2 and Omicron BA.1 respectively) compared to those seen for Fc effector functions and binding which ranged from 1 to 3 fold (Figure 1 C, F, I). Notably, Omicron infection elicited robust and similar neutralization titers against itself regardless of vaccination status.

## Discussion

While the neutralization resistance of Omicron is well-defined, here we assess how effectively Omicron-elicited antibodies target D614G and other VOCs. We also measured neutralization activity against BA.2, which now accounts for most infections in South Africa, and is increasing globally. We show that in previously unvaccinated individuals, Omicron-triggered antibodies bind and perform Fc effector function with only slight loss against VOCs. However, with the exception of Omicron BA.2 which showed comparatively modest decreases, neutralization was significantly compromised against VOCs, indicating limited neutralization cross-reactivity of antibodies elicited by Omicron. In contrast, vaccinated individuals who subsequently became infected with Omicron, showed greatly improved cross-reactivity with high titers against Omicron BA.1, BA.2, D614G (one amino acid different from the vaccine spike), Beta, Delta and C.1.2.

We and others have shown that Fc effector function is largely preserved against VOCs in both convalescent and vaccine-elicited plasma (Kaplonek et al., 2022; Richardson et al., 2022). Also, as with neutralization, we have shown that Fc effector function triggered by Beta is more cross-reactive compared to antibodies elicited by D614G, indicating that the spike sequence of the eliciting immunogen impacts the extent of ADCC cross-reactivity (Moyo-Gwete et al., 2021; Richardson et al., 2022). Here, we show that Omicron infection similarly triggers differential ADCC cross-reactivity, with significantly decreased activity against D614G and Beta, but not Delta. This observation extended to vaccinated individuals, where ADCC was still significantly poorer against Beta. This differential targeting of ADCC-mediating antibodies indicates that they may preferentially bind sites that differ between Omicron and other VOCs. Alternatively, different VOCs may trigger antibodies with varied glycosylations and isotype, both of which modulate Fc effector function (Jennewein and Alter, 2017).

This differential immune imprinting by VOCs was also confirmed for neutralization in this study in both unvaccinated and vaccinated Omicron-infected individuals. The highest titers in unvaccinated individuals were to the infection-matched Omicron. In contrast, in previously vaccinated individuals with breakthrough Omicron infections, high titers were observed against both D614G and Omicron; sequences that match the vaccine and infecting spikes to which these donors have been exposed. This is consistent with our studies of immune responses triggered by D614G, Beta or Delta (Keeton et al., 2021; Moyo-Gwete et al., 2021; Wibmer et al., 2021) which resulted in distinct patterns of cross-reactivity compared to those observed here after Omicron infection. Together these data indicate that the sequence of the infecting spike impacts the quality of neutralization, suggesting imprinting of the immune response (Keeton et al., 2021).

We and others have shown that humoral function is significantly boosted in individuals with breakthrough infections after vaccination (Kitchin et al., 2022; Suryawanshi et al., 2022; Walls et al., 2022). This study confirms this is also true of Omicron breakthrough infections, with a 153-fold increase in titers to D614G in vaccinated compared to unvaccinated individuals, consistent with other studies (Khan et al., 2022; Suryawanshi et al., 2022; Zhou et al., 2022). We also extend our previous study, where we showed ADCC was boosted by breakthrough infection, to include ADCP, confirming that this applies to other Fc effector functions (Kitchin et al., 2022).

In the absence of vaccination, Omicron-elicited humoral responses, while potent against the matched Omicron spike, show significantly less activity against VOCs. Thus, while highly immunogenic, Omicron does not elicit cross-neutralizing responses. This is consistent with a decrease in the ability of plasma from unvaccinated individuals to neutralize Delta compared to Omicron following Omicron infection (Khan et al., 2022). This may result in risk of reinfection in this unvaccinated group with other variants that continued to circulate and evolve in South Africa at the time of this study, including Beta, Delta and C.1.2. However, against Omicron BA.2, we noted only modest decreases in neutralizing titer relative to Omicron BA.1 in this cohort. This is in line with a study that shows 3-fold loss in activity in Omicron BA.1 infected hamsters against Omicron BA.2 (Yamasoba et al., 2022). This indicates that despite a number of differences between the sub-lineages, these changes do not seem to greatly alter the capacity of Omicron BA.1 antibodies to neutralize Omicron BA.2.

Our data also have implications for the design of second-generation vaccines based on Omicron, suggesting these may not trigger cross-reactive *de novo* responses in SARS-CoV-2 naive individuals. This is supported by immunogenicity studies, where Delta-infected mice elicited broadly protective antibodies, but Omicron-infected mice failed to mount responses against other VOCs (Suryawanshi et al., 2022). In addition, immunization of mice with a receptor binding domain (RBD)-based Omicron mRNA vaccine only elicited strain-specific neutralization (Lee et al., 2022). In support of this study and others (Hawman et al., 2022), our data also suggests that Omicron is highly immunogenic, eliciting comparable neutralization titers irrespective of vaccination status but through a largely strain-specific response. Given the significantly higher titers we see against Omicron in vaccinated individuals, Omicron boosters may be effective in seropositive individuals, a group that exceeds 70% in South Africa (Madhi et al., 2022). However, in a comparison of mRNA-1273 vaccinated rhesus macaques boosted with either mRNA-Omicron or mRNA-1273, Omicron boosted animals showed lower titers than those with a homologous mRNA-1273 boost (Gagne et al., 2022). Overall, these data suggest that boosting individuals with or without immunity with vaccines specific for Omicron is unlikely to be superior to existing regimens.

## Limitations of study

Our study is limited by the fact that we cannot rule out prior asymptomatic infection which could alter the quality of humoral responses. Sampling for this study was limited to blood, and occurred early in infection when antibodies may not have reached peak cross-reactivity. Furthermore, viral sequences confirming Omicron BA.1 infection are available only for a subset of samples. However, Omicron BA.1 overwhelmingly dominated infections during the wave in which these individuals were tested (Viana et al., 2022). Further, the median age of individuals in this study is high (58 years) and may contribute to the compromised cross-reactivity in unvaccinated individuals. Given the high prevalence of global seropositivity, through vaccination or previous infection, the ability to measure the response to Omicron infection in naive samples is limited. As such our study offers valuable insights into the value of Omicron as an immunogen and the risk of reinfection in unvaccinated individuals.

## Data Availability

All data produced in the present study are available upon reasonable request to the authors

## Acknowledgements

We acknowledge the participants who volunteered for this study and thank W van Hougenhouck-Tulleken and R Ramlall for database support and participant recruitment and D Amoako and J Giandhari for sequencing. The parental soluble spike was provided by J McLellan (University of Texas) and parental pseudovirus plasmids by Drs E Landais and D Sok (IAVI). PLM is supported by the South African Research Chairs Initiative of the Department of Science and Innovation and National Research Foundation of South Africa, the SA Medical Research Council SHIP program, the Centre for the AIDS Programme of Research in South Africa (CAPRISA). We acknowledge funding from the Bill and Melinda Gates Foundation, through the Global Immunology and Immune Sequencing for Epidemic Response (GIISER) program. SIR is a L’Oreal/UNESCO Women in Science South Africa Young Talents awardee.

## Author contributions

S.I.R designed the study, performed experiments, analyzed the data and wrote the manuscript. N.P.M and H.S performed Fc experiments and analyzed data. V.S.M, N.M and T.M performed neutralization assays. B.E.L, B.O. and F.A. produced spikes and variant plasmids. F.A and Z.M performed ELISA assays supervised by T.M.G. M.A. vdM processed samples which were recruited by Z. dB., T.R d. V., A.B. and G.vdB. V.U, T.R. and M.T.B established the Pretoria COVID-19 study which provided participant samples from the Tshwane District Hospital. A.S, A.M, H.T., M.V., and TdO performed SARS-CoV-2 sequencing. PLM conceptualized the study and wrote the manuscript.

## STAR Methods RESOURCE AVAILABILITY

### Lead Contact

Further information and reasonable requests for resources and reagents should be directed to and will be fulfilled by the lead contact, Penny Moore.

### Materials availability

Materials will be made by request to Penny Moore.

### Data and code availability

- All data reported in this paper will be shared by the lead contact upon request.
- This paper does not report original code.
- Any additional information required to reanalyze the data reported in this paper is available from the Lead Contact upon request.

## EXPERIMENTAL MODEL AND SUBJECT DETAILS

### Human Subjects

Samples infected in the fourth COVID-19 wave of infection in South Africa were collected from participants enrolled to the Pretoria COVID-19 study cohort. Participants were admitted to Tshwane District Hospital (Pretoria, South Africa) with mild to severe (WHO severity scale 3-6) PCR confirmed SARS-CoV-2 infection between 25 November 2021-20 December 2021 (Table S1). Ethics approval was received from the University of Pretoria, Human Research Ethics Committee (Medical) (247/2020). Second wave plasma samples were obtained from hospitalized COVID-19 patients with moderate disease (WHO scale 4-5) admitted to Groote Schuur Hospital cohort, Cape Town from December 2020 – January 2021. This study received ethics approval from the Human Research Ethics Committee of the Faculty of Health Sciences, University of Cape Town (R021/2020). Wave 3 samples were collected from hospitalized COVID-19 patients with moderate disease (WHO scale 4-5) from Steve Biko Academic Hospital and Groote Schuur Hospital in July 2021. All patients had PCR confirmed SARS-CoV-2 infection before blood collection which was done a median of 4 days post positive PCR test. Healthy donors who were confirmed to be SARS-CoV-2 infection naïve and were vaccinated with either one dose of Janssen/Johnson and Johnson (Ad26.COV2.S) vaccine during the Sisonke Trial or two doses of BNT162b2 were obtained 2 months after full vaccination. Ethics approval for the use of these samples were obtained from the Human Research Ethics Committee of the Faculty of Health Sciences, University of the Witwatersrand (M210465). Written informed consent was obtained from all participants.

### Cell lines

Human embryo kidney HEK293T cells were cultured at 37°C, 5% CO2, in DMEM containing 10% heat-inactivated fetal bovine serum (Gibco BRL Life Technologies) and supplemented with 50 μg/ml gentamicin (Sigma). Cells were disrupted at confluence with 0.25% trypsin in 1 mM EDTA (Sigma) every 48–72 hours. HEK293T/ACE2.MF cells were maintained in the same way as HEK293T cells but were supplemented with 3 μg/ml puromycin for selection of stably transduced cells. HEK293F suspension cells were cultured in 293 Freestyle media (Gibco BRL Life Technologies) and cultured in a shaking incubator at 37°C, 5% CO2, 70% humidity at 125rpm maintained between 0.2 and 0.5 million cells/ml. Jurkat-Lucia™ NFAT-CD16 cells were maintained in IMDM media with 10% heat-inactivated fetal bovine serum (Gibco, Gaithersburg, MD), 1% Penicillin Streptomycin (Gibco, Gaithersburg, MD) and 10 μg/ml of Blasticidin and 100 μg/ml of Zeocin was added to the growth medium every other passage. THP-1 cells were used for the ADCP assay and obtained from the AIDS Reagent Program, Division of AIDS, NIAID, NIH contributed by Dr. Li Wu and Vineet N. KewalRamani. Cells were cultured at 37°C, 5% CO2 in RPMI containing 10% heat-inactivated fetal bovine serum (Gibco, Gaithersburg, MD) with 1% Penicillin Streptomycin (Gibco, Gaithersburg, MD) and 2-mercaptoethanol to a final concentration of 0.05 mM and not allowed to exceed 4 × 10^5^ cells/ml to prevent differentiation.

## METHOD DETAILS

### SARS-CoV-2 spike genome sequencing

Sequencing of the spike was performed as previously described (Tegally et al., 2021) using swabs obtained from Tshwane District Hospital patients of which 6 were available and confirmed to be Omicron BA.1 (Table S1). RNA sequencing was performed as previously published. Briefly, extracted RNA was used to synthesize cDNA using the Superscript IV First Strand synthesis system (Life Technologies, Carlsbad, CA) and random hexamer primers. SARS-CoV-2 whole genome amplification was performed by multiplex PCR using primers designed on Primal Scheme (http://primal.zibraproject.org/) to generate 400 bp amplicons with a 70 bp overlap covering the SARS-CoV-2 genome. Phylogenetic clade classification of the genomes in this study consisted of analyzing them against a global reference dataset using a custom pipeline based on a local version of NextStrain (https://github.com/nextstrain/ncov) (Hadfield et al., 2018).

### SARS-CoV-2 antigens

For ELISA and ADCP assays, SARS-CoV-2 original and Beta variant full spike (L18F, D80A, D215G, K417N, E484K, N501Y, D614G, A701V, 242-244 del), Delta (T19R, 156-157del, R158G, L452R, T478K, D614G, P681R and D950N) and Omicron BA.1 (A67V, Δ69-70, T95I, G142D, Δ143-145, Δ211, L212I, 214EPE, G339D, S371L, S373P, S375F, K417N, N440K, G446S, S477N, T478K, E484A, Q493R, G496S, Q498R, N501Y, Y505H, T547K, D614G, H655Y, N679K, P681H, N764K, D796Y, N856K, Q954H, N969K, L981F) proteins were expressed in Human Embryonic Kidney (HEK) 293F suspension cells by transfecting the cells with the respective expression plasmid. After incubating for six days at 37 °C, 70% humidity and 10% CO_2_, proteins were first purified using a nickel resin followed by size-exclusion chromatography. Relevant fractions were collected and frozen at -80 °C until use.

### SARS-CoV-2 Spike Enzyme-linked immunosorbent assay (ELISA)

Two μg/ml of spike protein (D614G, Beta, Delta or Omicron) was used to coat 96-well, high-binding plates and incubated overnight at 4 °C. The plates were incubated in a blocking buffer consisting of 5% skimmed milk powder, 0.05% Tween 20, 1x PBS. Plasma samples were diluted to 1:100 starting dilution in a blocking buffer and added to the plates. IgG secondary antibody was diluted to 1:3000 in blocking buffer and added to the plates followed by TMB substrate (Thermofisher Scientific). Upon stopping the reaction with 1 M H_2_SO_4_, absorbance was measured at a 450nm wavelength. In all instances, mAbs CR3022 and BD23 were used as positive controls and Palivizumab was used as a negative control.

### Spike plasmid and Lentiviral Pseudovirus Production

The SARS-CoV-2 Wuhan-1 spike, cloned into pCDNA3.1 was mutated using the QuikChange Lightning Site-Directed Mutagenesis kit (Agilent Technologies) and NEBuilder HiFi DNA Assembly Master Mix (NEB) to include D614G (original) or lineage defining mutations for Beta (L18F, D80A, D215G, 242-244del, K417N, E484K, N501Y, D614G and A701V), Delta (T19R, 156-157del, R158G, L452R, T478K, D614G, P681R and D950N), C.1.2. (P9L, P25L, C136F, Δ144, R190S, D215G, Δ242-243, Y449H, E484K, N501Y, L585F, D614G, H655Y, N679K, T716I, T859N), Omicron BA.1 (A67V, Δ69-70, T95I, G142D, Δ143-145, Δ211, L212I, 214EPE, G339D, S371L, S373P, S375F, K417N, N440K, G446S, S477N, T478K, E484A, Q493R, G496S, Q498R, N501Y, Y505H, T547K, D614G, H655Y, N679K, P681H, N764K, D796Y, N856K, Q954H, N969K, L981F) or Omicron BA.2 (T19I, L24S, 25-27del, G142D, V213G, G339D, S371F, S373P, S375F, T376A, D405N, R408S,K417N,N440K, S477N, T478K, E484A, Q493R, Q498R, N501Y, Y505H, D614G, H655Y, N679K, P681H, N764K, D796Y, Q954H, N969K).

Pseudotyped lentiviruses were prepared by co-transfecting HEK293T cell line with the SARS-CoV-2 ancestral variant spike (D614G), Beta, Delta, C.1.2, Omicron BA.1 or Omicron BA.2 spike plasmids in conjunction with a firefly luciferase encoding lentivirus backbone (HIV-1 pNL4.luc) plasmid as previously described (Wibmer et al., 2021). Culture supernatants were clarified of cells by a 0.45-μM filter and stored at -70 °C. Other pcDNA plasmids were used for the ADCC assay.

### Pseudovirus neutralization assay

For the neutralization assay, plasma samples were heat-inactivated and clarified by centrifugation. Heat-inactivated plasma samples from vaccine recipients were incubated with the SARS-CoV-2 pseudotyped virus for 1 hour at 37°C, 5% CO2. Subsequently, 1×10^4^ HEK293T cells engineered to over-express ACE-2 (293T/ACE2.MF)(kindly provided by M. Farzan (Scripps Research)) were added and incubated at 37°C, 5% CO_2_ for 72 hours upon which the luminescence of the luciferase gene was measured. Titers were calculated as the reciprocal plasma dilution (ID_50_) causing 50% reduction of relative light units. CB6 and CA1 was used as positive controls for D614G, Beta and Delta. 084-7D, a mAb targeting K417N was used as a positive control for Omicron and Beta.

### Antibody-dependent cellular phagocytosis (ADCP) assay

Avitagged SARS-CoV-2 spikes were biotinylated using the BirA biotin-protein ligase standard reaction kit (Avidity, LLC) and coated onto fluorescent neutravidin beads as previously described (Ackerman et al., 2011). Briefly, beads were incubated for two hours with monoclonal antibodies at a starting concentration of 20 μg/ml or plasma at a single 1 in 100 dilution. Opsonized beads were incubated with the monocytic THP-1 cell line overnight, fixed and interrogated on the FACSAria II. Phagocytosis score was calculated as the percentage of THP-1 cells that engulfed fluorescent beads multiplied by the geometric mean fluorescence intensity of the population less the no antibody control. For this and all subsequent Fc effector assays, pooled plasma from 5 PCR-confirmed SARS-CoV-2 infected individuals and CR3022 were used as positive controls and plasma from 5 pre-pandemic healthy controls and Palivizumab were used as negative controls. 084-7D was used as a positive control for Omicron BA.1 and Beta. In addition samples both waves were run head-to-head in the same experiment. ADCP scores for different spikes were normalised to each other and between runs using CR3022.

### Antibody-dependent cellular cytotoxicity (ADCC) assay

The ability of plasma antibodies to cross-link and signal through FcγRIIIa (CD16) and spike expressing cells or SARS-CoV-2 protein was measured as a proxy for ADCC. For spike assays, HEK293T cells were transfected with 5μg of SARS-CoV-2 spike plasmids using PEI-MAX 40,000 (Polysciences) and incubated for 2 days at 37°C. Expression of spike was confirmed by differential binding of CR3022 and P2B-2F6 and their detection by anti-IgG APC staining measured by flow cytometry. Subsequently, 1×10^5^ spike transfected cells per well were incubated with heat inactivated plasma (1:100 final dilution) or monoclonal antibodies (final concentration of 100 μg/ml) in RPMI 1640 media supplemented with 10% FBS 1% Pen/Strep (Gibco, Gaithersburg, MD) for 1 hour at 37°C. Jurkat-Lucia™ NFAT-CD16 cells (Invivogen) (2×10^5^ cells/well and 1×10^5^ cells/well for spike and other protein respectively) were added and incubated for 24 hours at 37°C, 5% CO_2_. Twenty μl of supernatant was then transferred to a white 96-well plate with 50 μl of reconstituted QUANTI-Luc secreted luciferase and read immediately on a Victor 3 luminometer with 1s integration time. Relative light units (RLU) of a no antibody control was subtracted as background. Palivizumab was used as a negative control, while CR3022 was used as a positive control, and P2B-2F6 to differentiate the Beta from the D614G variant. 084-7D was used as a positive control for Omicron BA.1 and Beta. To induce the transgene 1x cell stimulation cocktail (Thermofisher Scientific, Oslo, Norway) and 2 μg/ml ionomycin in R10 was added as a positive control to confirm sufficient expression of the Fc receptor. RLUs for spikes were normalised to each other and between runs using CR3022. All samples were run head to head in the same experiment as were all variants tested.

## QUANTIFICATION AND STATISTICAL ANALYSIS

Analyses were performed in Prism (v9; GraphPad Software Inc, San Diego, CA, USA). Non-parametric tests were used for all comparisons. The Mann-Whitney and Wilcoxon tests were used for unmatched and paired samples, respectively. The Friedman test with Dunns correction for multiple comparisons was used for matched comparisons across variants. All correlations reported are non-parametric Spearman’s correlations. *P* values less than 0.05 were considered to be statistically significant.

## References

Ackerman, M.E., Moldt, B., Wyatt, R.T., Dugast, A.-S., McAndrew, E., Tsoukas, S., Jost, S., Berger, C.T., Sciaranghella, G., Liu, Q., et al. (2011). A robust, high-throughput assay to determine the phagocytic activity of clinical antibody samples. J. Immunol. Methods 366, 8–19.

Bartsch, Y., Tong, X., Kang, J., Avendaño, M.J., Serrano, E.F., García-Salum, T., Pardo-Roa, C., Riquelme, A., Medina, R.A., and Alter, G. (2021). Preserved Omicron Spike specific antibody binding and Fc-recognition across COVID-19 vaccine platforms. MedRxiv 2021.12.24.21268378.

Carreño, J.M., Alshammary, H., Tcheou, J., Singh, G., Raskin, A., Kawabata, H., Sominsky, L., Clark, J., Adelsberg, D.C., Bielak, D., et al. (2021). Activity of convalescent and vaccine serum against SARS-CoV-2 Omicron. Nature.

Cele, S., Jackson, L., Khoury, D.S., Khan, K., Moyo-Gwete, T., Tegally, H., San, J.E., Cromer, D., Scheepers, C., Amoako, D.G., et al. (2021). Omicron extensively but incompletely escapes Pfizer BNT162b2 neutralization. Nature.

Collie, S., Champion, J., Moultrie, H., Bekker, L.-G., and Gray, G. (2022). Effectiveness of BNT162b2 Vaccine against Omicron Variant in South Africa. N. Engl. J. Med. 386, 494–496.

Gagne, M., Moliva, J.I., Foulds, K.E., Andrew, S.F., Flynn, B.J., Werner, A.P., Wagner, D.A., Teng, I.-T., Lin, B.C., Moore, C., et al. (2022). mRNA-1273 or mRNA-Omicron boost in vaccinated macaques elicits comparable B cell expansion, neutralizing antibodies and protection against Omicron. BioRxiv 2022.02.03.479037.

Gao, Y., Cai, C., Grifoni, A., Müller, T.R., Niessl, J., Olofsson, A., Humbert, M., Hansson, L., Österborg, A., Bergman, P., et al. (2022). Ancestral SARS-CoV-2-specific T cells cross-recognize the Omicron variant. Nat. Med.

Garcia-Beltran, W.F., St. Denis, K.J., Hoelzemer, A., Lam, E.C., Nitido, A.D., Sheehan, M.L., Berrios, C., Ofoman, O., Chang, C.C., Hauser, B.M., et al. (2022). mRNA-based COVID-19 vaccine boosters induce neutralizing immunity against SARS-CoV-2 Omicron variant. Cell 185, 457–466.e4.

Hadfield, J., Megill, C., Bell, S.M., Huddleston, J., Potter, B., Callender, C., Sagulenko, P., Bedford, T., and Neher, R.A. (2018). Nextstrain: real-time tracking of pathogen evolution. Bioinformatics 34, 4121–4123.

Hawman, D.W., Meade-White, K., Clancy, C., Archer, J., Hinkley, T., Leventhal, S.S., Rao, D., Stamper, A., Lewis, M., Rosenke, R., et al. (2022). Replicating RNA platform enables rapid response to the SARS-CoV-2 Omicron variant and elicits enhanced protection in naïve hamsters compared to ancestral vaccine. BioRxiv 2022.01.31.478520.

Jennewein, M.F., and Alter, G. (2017). The Immunoregulatory Roles of Antibody Glycosylation. Trends Immunol. 38, 358–372.

Kaplonek, P., Fischinger, S., Cizmeci, D., Bartsch, Y.C., Kang, J., Burke, J.S., Shin, S.A., Dayal, D., Martin, P., Mann, C., et al. (2022). mRNA-1273 vaccine-induced antibodies maintain Fc effector functions across SARS-CoV-2 variants of concern. Immunity S1074-7613(22)00030-9.

Keeton, R., Richardson, S.I., Moyo-Gwete, T., Hermanus, T., Tincho, M.B., Benede, N., Manamela, N.P., Baguma, R., Makhado, Z., Ngomti, A., et al. (2021). Prior infection with SARS-CoV-2 boosts and broadens Ad26.COV2.S immunogenicity in a variant-dependent manner. Cell Host Microbe 29, 1611–1619.e5.

Keeton, R., Tincho, M.B., Ngomti, A., Baguma, R., Benede, N., Suzuki, A., Khan, K., Cele, S., Bernstein, M., Karim, F., et al. (2022). T cell responses to SARS-CoV-2 spike cross-recognize Omicron. Nature.

Khan, K., Karim, F., Cele, S., San, J.E., Lustig, G., Tegally, H., Rosenberg, Y., Bernstein, M., Ganga, Y., Jule, Z., et al. (2022). Omicron infection of vaccinated individuals enhances neutralizing immunity against the Delta variant. MedRxiv 2021.12.27.21268439.

Kitchin, D., Richardson, S.I., van der Mescht, M.A., Motlou, T., Mzindle, N., Moyo-Gwete, T., Makhado, Z., Ayres, F., Manamela, N.P., Spencer, H., et al. (2022). Ad26.COV2.S breakthrough infections induce high titers of neutralizing antibodies against Omicron and other SARS-CoV-2 variants of concern. Cell Reports Med. 100535.

Lee, I.-J., Sun, C.-P., Wu, P.-Y., Lan, Y.-H., Wang, I.-H., Liu, W.-C., Tseng, S.-C., Tsung, S.-I., Chou, Y.-C., Kumari, M., et al. (2022). Omicron-specific mRNA vaccine induced potent neutralizing antibody against Omicron but not other SARS-CoV-2 variants. BioRxiv 2022.01.31.478406.

Madhi, S.A., Kwatra, G., Myers, J.E., Jassat, W., Dhar, N., Mukendi, C.K., Nana, A.J., Blumberg, L., Welch, R., Ngorima-Mabhena, N., et al. (2022). South African Population Immunity and Severe Covid-19 with Omicron Variant. MedRxiv 2021.12.20.21268096.

Moyo-Gwete, T., Madzivhandila, M., Makhado, Z., Ayres, F., Mhlanga, D., Oosthuysen, B., Lambson, B.E., Kgagudi, P., Tegally, H., Iranzadeh, A., et al. (2021). Cross-Reactive Neutralizing Antibody Responses Elicited by SARS-CoV-2 501Y.V2 (B.1.351). N. Engl. J. Med. 384, 2161–2163.

Richardson, S.I., Manamela, N.P., Motsoeneng, B.M., Kaldine, H., Ayres, F., Makhado, Z., Mennen, M., Skelem, S., Williams, N., Sullivan, N.J., et al. (2022). SARS-CoV-2 Beta and Delta variants trigger Fc effector function with increased cross-reactivity. Cell Reports Med. 100510.

Scheepers, C., Everatt, J., Amoako, D.G., Tegally, H., Wibmer, C.K., Mnguni, A., Ismail, A., Mahlangu, B., Lambson, B.E., Richardson, S.I., et al. (2021). Emergence and phenotypic characterization of C.1.2, a globally detected lineage that rapidly accumulated mutations of concern. MedRxiv 2021.08.20.21262342.

Schmidt, F., Muecksch, F., Weisblum, Y., Da Silva, J., Bednarski, E., Cho, A., Wang, Z., Gaebler, C., Caskey, M., Nussenzweig, M.C., et al. (2021). Plasma Neutralization of the SARS-CoV-2 Omicron Variant. N. Engl. J. Med.

Sievers, B.L., Chakraborty, S., Xue, Y., Gelbart, T., Gonzalez, J.C., Cassidy, A.G., Golan, Y., Prahl, M., Gaw, S.L., Arunachalam, P.S., et al. (2022). Antibodies elicited by SARS-CoV-2 infection or mRNA vaccines have reduced neutralizing activity against Beta and Omicron pseudoviruses. Sci. Transl. Med. 0, eabn7842.

Suryawanshi, R.K., Chen, I.P., Ma, T., Syed, A.M., Simoneau, C.R., Ciling, A., Khalid, M.M., Sreekumar, B., Chen, P.-Y., George, A.F., et al. (2022). Limited cross-variant immunity after infection with the SARS-CoV-2 Omicron variant without vaccination. MedRxiv 2022.01.13.22269243.

Tarke, A., Coelho, C.H., Zhang, Z., Dan, J.M., Yu, E.D., Methot, N., Bloom, N.I., Goodwin, B., Phillips, E., Mallal, S., et al. (2022). SARS-CoV-2 vaccination induces immunological T cell memory able to cross-recognize variants from Alpha to Omicron. Cell.

Tegally, H., Wilkinson, E., Giovanetti, M., Iranzadeh, A., Fonseca, V., Giandhari, J., Doolabh, D., Pillay, S., San, E.J., Msomi, N., et al. (2021). Detection of a SARS-CoV-2 variant of concern in South Africa. Nature 592, 438–443.

Viana, R., Moyo, S., Amoako, D.G., Tegally, H., Scheepers, C., Althaus, C.L., Anyaneji, U.J., Bester, P.A., Boni, M.F., Chand, M., et al. (2022). Rapid epidemic expansion of the SARS-CoV-2 Omicron variant in southern Africa. Nature.

Walls, A.C., Sprouse, K.R., Bowen, J.E., Joshi, A., Franko, N., Navarro, M.J., Stewart, C., Cameroni, E., McCallum, M., Goecker, E.A., et al. (2022). SARS-CoV-2 breakthrough infections elicit potent, broad, and durable neutralizing antibody responses. Cell.

Wibmer, C.K., Ayres, F., Hermanus, T., Madzivhandila, M., Kgagudi, P., Oosthuysen, B., Lambson, B.E., de Oliveira, T., Vermeulen, M., van der Berg, K., et al. (2021). SARS-CoV-2 501Y.V2 escapes neutralization by South African COVID-19 donor plasma. Nat. Med.

Yamasoba, D., Kimura, I., Nasser, H., Morioka, Y., Nao, N., Ito, J., Uriu, K., Tsuda, M., Zahradnik, J., Shirakawa, K., et al. (2022). Virological characteristics of SARS-CoV-2 BA.2 variant. BioRxiv 2022.02.14.480335.

Yu, J., Tostanoski, L.H., Peter, L., Mercado, N.B., McMahan, K., Mahrokhian, S.H., Nkolola, J.P., Liu, J., Li, Z., Chandrashekar, A., et al. (2020). DNA vaccine protection against SARS-CoV-2 in rhesus macaques. Science (80-.). 369, 806–811.

Yu, J., Collier, A.-R.Y., Rowe, M., Mardas, F., Ventura, J.D., Wan, H., Miller, J., Powers, O., Chung, B., Siamatu, M., et al. (2022). Comparable Neutralization of the SARS-CoV-2 Omicron BA.1 and BA.2 Variants. MedRxiv Prepr. Serv. Heal. Sci. 2022.02.06.22270533.

Zhou, R., To, K.K.-W., Peng, Q., Chan, J.M.-C., Huang, H., Yang, D., Lam, B.H.-S., Chuang, V.W.-M., Cai, J.-P., Liu, N., et al. (2022). Vaccine-breakthrough infection by the SARS-CoV-2 omicron variant elicits broadly cross-reactive immune responses. Clin. Transl. Med. 12, e720–e720.

Zohar, T., Loos, C., Fischinger, S., Atyeo, C., Wang, C., Slein, M.D., Burke, J., Yu, J., Feldman, J., Hauser, B.M., et al. (2020). Compromised Humoral Functional Evolution Tracks with SARS-CoV-2 Mortality. Cell 183, 1508–1519.e12.

