## Supplemental data for "SARS-CoV-2 Omicron triggers cross-reactive neutralization and Fc effector functions in previously vaccinated, but not unvaccinated individuals"

### Table S1: Demographic and clinical description of the cohort, Related to STAR Methods

| Study number | Age | Gender | Days sampled post infection | Vaccination status | Days since vaccination | Sequence | Severity |
|---|---|---|---|---|---|---|---|
| COV132 | 36-40 | Male | 9 | Unvaccinated | - | Not sequenced | Severe |
| COV140 | 26-30 | Female | 1 | Unvaccinated | - | Not sequenced | Mild |
| COV145 | 16-20 | Female | 3 | Unvaccinated | - | Omicron | Mild |
| COV149 | 60-65 | Female | 3 | Unvaccinated | - | Not sequenced | Mild |
| COV157 | 66-70 | Male | 4 | Unvaccinated | - | Omicron | Moderate |
| COV158 | 66-70 | Female | 6 | Unvaccinated | - | Not sequenced | Moderate |
| COV159 | 20-25 | Male | 6 | Unvaccinated | - | Not sequenced | Mild |
| COV162 | 56-60 | Male | 5 | Unvaccinated | - | Not sequenced | Moderate |
| COV165 | 60-65 | Female | 4 | Unvaccinated | - | Not sequenced | Moderate |
| COV166 | 56-60 | Male | 4 | Unvaccinated | - | Not sequenced | Severe |
| COV167 | 60-65 | Female | 8 | Unvaccinated | - | Not sequenced | Severe |
| COV176 | 70-75 | Male | 2 | Unvaccinated | - | Not sequenced | Moderate |
| COV178 | 60-65 | Female | 2 | Unvaccinated | - | Omicron | Moderate |
| COV183 | 26-30 | Male | 2 | Unvaccinated | - | Not sequenced | Mild |
| COV184 | 56-60 | Male | 1 | Unvaccinated | - | Not sequenced | Moderate |
| COV185 | 46-50 | Female | 1 | Unvaccinated | - | Not sequenced | Moderate |
| COV186 | 56-60 | Male | 10 | Unvaccinated | - | Omicron | Moderate |
| COV187 | 30-35 | Female | 3 | Unvaccinated | - | Not sequenced | Mild |
| COV189 | 46-50 | Male | 5 | Unvaccinated | - | Not sequenced | Moderate |
| COV190 | 30-35 | Female | 4 | Unvaccinated | - | Omicron | Mild |
| COV151 | 46-50 | Male | 1 | One dose Ad26.CoV2.S | 56 | Omicron | Severe |
| COV182 | 36-40 | Male | 3 | One dose Ad26.CoV2.S | Unknown | Not sequenced | Mild |
| COV131 | 56-60 | Female | 4 | Two doses BNT162b2 | 109 | Not sequenced | Severe |
| COV135 | 80-85 | Male | 5 | Two doses BNT162b2 | 72 | Omicron | Mild |
| COV138 | 66-70 | Male | 5 | Two doses BNT162b2 | Unknown | Not sequenced | Moderate |
| COV161 | 60-65 | Female | 5 | Two doses BNT162b2 | 112 | Not sequenced | Mild |
| COV181 | 70-75 | Male | 4 | Two doses BNT162b2 | 163 | Not sequenced | Severe |

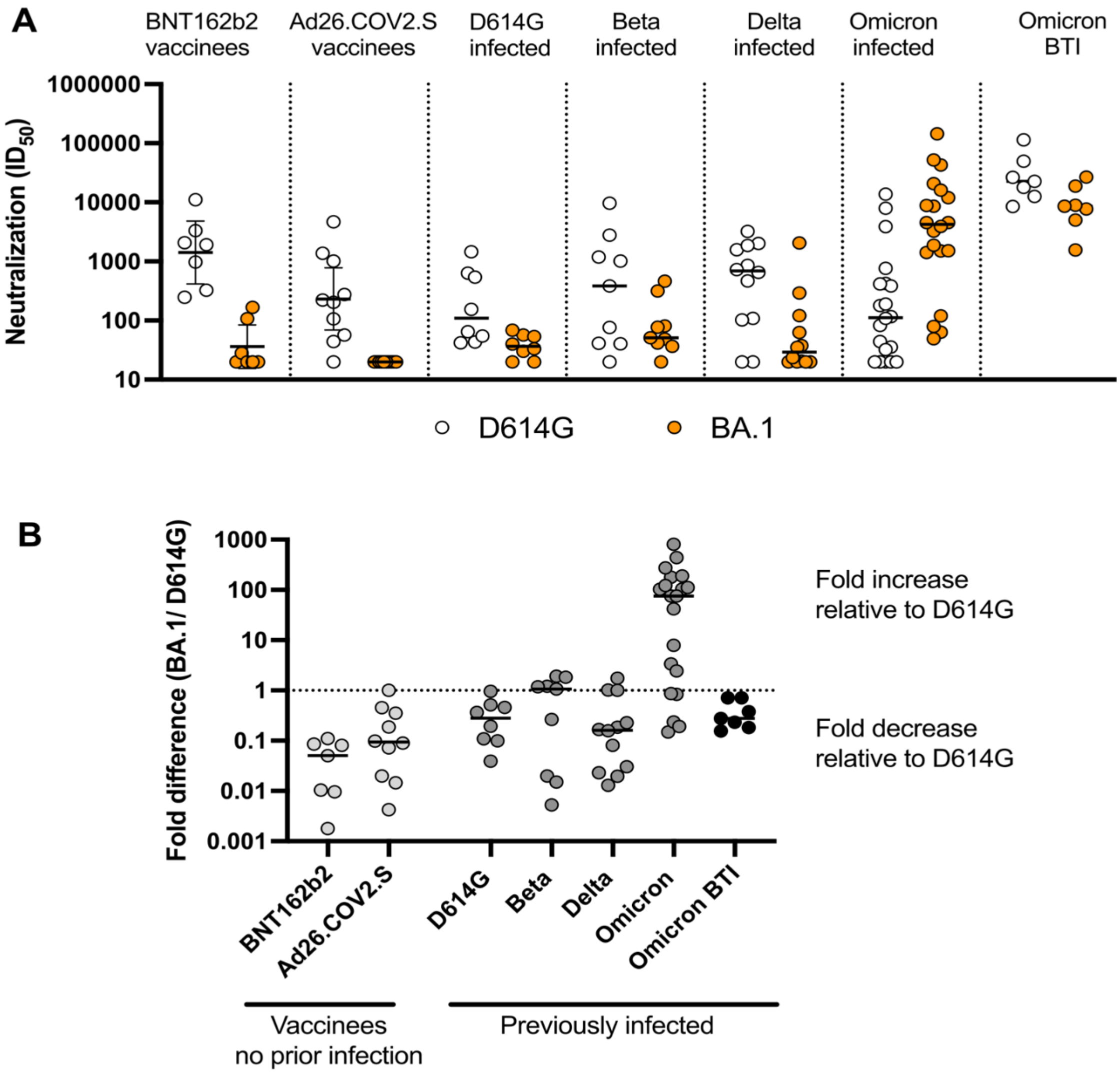


**Figure S1: Omicron BA.1 neutralization titers are substantially boosted in response to Omicron infection compared to infection with other VOCs or vaccination, Related to Figure 2**

(A) Neutralization $ID_{50}$ titers from vaccinees two months post two-doses of BNT162b2 or one-dose of Ad26.COV2.S, individuals acutely infected with D614G, Beta, Delta or Omicron or individuals who were vaccinated with either BNT162b2 or Ad26.COV2.S and then infected with Omicron (BTI = breakthrough infection) against D614G (white) and Omicron BA.1 (orange) SARS-CoV-2 variants. Lines indicate geometric mean titer (GMT). (B) Fold difference of neutralization titer against Omicron BA.1 relative to D614G for each individual is shown with median fold difference indicated by the bold line. The dotted line indicates no fold difference.
